# Namilumab or infliximab compared to standard of care in hospitalised patients with COVID-19 (CATALYST): a phase 2 randomised adaptive trial

**DOI:** 10.1101/2021.06.02.21258204

**Authors:** Benjamin A. Fisher, Tonny Veenith, Daniel Slade, Charlotte Gaskell, Matthew Rowland, Tony Whitehouse, James Scriven, Dhruv Parekh, Madhu S. Balasubramaniam, Graham Cooke, Nick Morley, Zoe Gabriel, Matthew P. Wise, Joanna Porter, Helen McShane, Ling-Pei Ho, Philip N. Newsome, Anna Rowe, Rowena Sharpe, David R. Thickett, Julian Bion, Simon Gates, Duncan Richards, Pamela Kearns, on behalf of CATALYST investigators

## Abstract

**Background:** Dysregulated inflammation is associated with poor outcomes in Coronavirus disease 2019 (COVID-19). We assessed the efficacy of namilumab, a granulocyte-macrophage colony-stimulating factor inhibitor and infliximab, a tumour necrosis factor inhibitor in hospitalised patients with COVID-19 in order to prioritise agents for phase 3 trials.

**Methods:** In this randomised, multi-arm, parallel group, open label, adaptive phase 2 proof-of-concept trial (CATALYST) we recruited hospitalised patients ≥ 16 years with COVID-19 pneumonia and C-reactive protein (CRP) ≥ 40mg/L in nine UK hospitals. Participants were randomly allocated with equal probability to usual care, or usual care plus a single 150mg intravenous dose of namilumab (150mg) or infliximab (5mg/kg). Randomisation was stratified for ward versus ICU. The primary endpoint was improvement in inflammation in intervention arms compared to control as measured by CRP over time, analysed using Bayesian multi-level models. ISRCTN registry number 40580903.

**Findings:** Between 15^th^ June 2020 and 18^th^ February 2021 we randomised 146 participants: 54 to usual care, 57 to namilumab and 35 to infliximab. The probabilities that namilumab and infliximab were superior to usual care in reducing CRP over time were 97% and 15% respectively. Consistent effects were seen in ward and ICU patients and aligned with clinical outcomes, such that the probability of discharge (WHO levels 1-3) at day 28 was 47% and 64% for ICU and ward patients on usual care, versus 66% and 77% for patients treated with namilumab. 134 adverse events occurred in 30/55 (54.5%) namilumab patients compared to 145 in 29/54 (53.7%) usual care patients. 102 events occurred in 20/29 (69.0%) infliximab patients versus 112 events in 17/34 (50.0%) usual care patients.

**Interpretation:** Namilumab, but not infliximab, demonstrated proof-of-concept evidence for reduction in inflammation in hospitalised patients with COVID-19 pneumonia which was consistent with secondary clinical outcomes. Namilumab should be prioritised for further investigation in COVID-19.

**Funding:** Medical Research Council.

## Introduction

Severe Coronavirus disease 2019 (COVID-19) is associated with high mortality and disability in survivors. An excessive and dysregulated immune response contributes to these poor outcomes, as evidenced by the ability of corticosteroids and IL-6 receptor blockade to reduce mortality in hospitalised patients requiring oxygen.^1,2^

Inflammatory monocytes/macrophages (IMM) appear central to this dysregulated response,^3^ resulting in disruption of pulmonary endothelial barrier integrity, microvascular thrombosis,^4^ and lung tissue damage.^5^ A genome-wide association study has identified the monocyte/macrophage chemotactic protein CCR2 as being associated with severe COVID-19.^6^ Transcriptomic analysis of blood, lung and bronchoalveolar fluid has revealed a predominance of activated IMM within the lung, alongside expression of pro-coagulant genes within alveolar macrophages.^7^ Notably, the aberrant expression of proliferation markers in blood monocytes correlates with severe disease,^8^ and likely reflects a pathological early release of monocytes from the bone marrow.^9^ IMM may be further activated and polarised to an inflammatory phenotype in severe disease by interaction with immune complexes containing hypoglycosylated anti-spike protein antibodies.^10^

IMM or their activity may be targeted therapeutically in a number of different ways. Given that trials with clinical outcomes require large numbers of patients to show effects, we designed a multi-arm proof of concept trial with a biomarker primary outcome to expedite decision-making on potentially effective therapeutic options for COVID-19. The aim was to provide early biological signals of efficacy to efficiently prioritise agents with the highest likelihood of success for study in established phase 3 platform trials.^11^ The first two agents studied were namilumab and infliximab.

Namilumab is an anti-granulocyte-macrophage colony stimulating factor (GM-CSF) monoclonal antibody with a good safety profile up to phase 2 that has been studied in inflammatory conditions such as rheumatoid arthritis. GM-CSF is a multifunctional cytokine that is a growth factor for granulocytes and monocytes and has an important role in immune responses. In particular, it drives the activation, maturation, survival and trafficking of monocyte-derived macrophages, and their polarisation towards a more inflammatory phenotype. Elevated GM-CSF levels are closely associated with disease severity in COVID-19,^12^ with GM-CSF-expressing T cells being clonally expanded in the lungs.^13^ Notably, GM-CSF may also enhance the pro-coagulant activities of macrophages,^14^ and blood clots are a recognised side effect of recombinant GM-CSF (sargramostim), suggesting that dysregulated GM-CSF expression may predispose to the microvascular thrombosis characteristic of COVID-19.^4^.

Infliximab is a widely used anti-tumour necrosis factor (TNF) monoclonal antibody. TNF is an important pro-inflammatory cytokine and its inhibition has shown efficacy in many chronic immune mediated inflammatory diseases (IMIDs). TNF inhibition reduces mortality and severity in several mouse models of viral respiratory infection.^15,16^ An IMM subset associated with severe COVID-19 shares transcriptional similarities to macrophages stimulated with both TNF and interferon gamma (IFNγ).^17^ Some data suggest that IMID patients who contract COVID-19 whilst treated with TNF inhibitors have better outcomes.^18^

We sought to provide early proof-of-concept signal in a randomised trial to efficiently prioritise these approaches for subsequent testing in larger trials powered for clinical outcomes.

## Methods

### Study design

The CATALYST trial is a randomised, open label, phase 2, multi-arm proof-of-concept trial.^11^ A placebo control was not included due to the operational difficulties imposed by the pandemic and the proposed multi-arm design and following advice from patient and public involvement. Participants were recruited from nine hospital sites in the UK (Queen Elizabeth Hospital Birmingham; Heartlands Hospital, Birmingham; John Radcliffe Hospital, Oxford; Royal Bolton Hospital, Bolton; Imperial St Mary’s Hospital, London; Royal Hallamshire Hospital, Sheffield; University Hospital of Wales, Cardiff; Good Hope Hospital, Birmingham and University College Hospital, London). The trial was approved by the East Midlands-Nottingham 2 Research Ethics Committee (20/EM/0115) and given national Urgent Public Health Status.

### Participants

Eligible patients were 16 years or over, with a clinical picture strongly suggestive of SARS-CoV-2 pneumonia (confirmed by chest X-ray or CT scan, with or without a positive reverse transcription-polymerase chain reaction (RT-PCR) assay), and with a C-Reactive Protein (CRP) ≥ 40 mg/L. The requirement for raised CRP replaced an inclusion criterion for low oxygenation status (oxygen saturation ≤94% while breathing ambient air or a ratio of the partial pressure of oxygen to the fraction of inspired oxygen ≤300 mmHg) early in the course of recruitment following a change in primary outcome (see below). Exclusion criteria included planned palliative care, pregnancy or breastfeeding, women of childbearing potential and non-vasectomised men who were unwilling to use effective contraception for the duration of the trial and throughout the drug-defined post-trial period, known HIV or chronic hepatitis B or C infection, concurrent immunosuppression with biological agents, a history of haematopoietic stem cell or solid organ transplant, known hypersensitivity to drug products or excipients, tuberculosis or other severe infections such as (non-SARS-CoV-2) sepsis, abscesses, and opportunistic infections requiring treatment, moderate or severe heart failure (NYHA class III/IV), or any other indication or medical history, that in the opinion of the patient’s local investigator, made the patient unsuitable for trial participation. Co-enrolment into other interventional trials was not permitted with the exception of the RECOVERY-Respiratory Support trial comparing continuous positive airway pressure or high flow nasal oxygen to standard care, as this met current UK guidance on mechanistic independence in co-enrolment.^19^

Written informed consent was obtained from all patients with capacity. If the patient lacked capacity, from severity of illness for example, informed consent was obtained from the patient’s personal legal representative or, if unavailable, a professional legal representative according to the requirements of the UK Health Research Authority. Patients with representative consent were re-consented as soon as possible after regaining capacity.

### Randomisation

Participants were randomly assigned with equal probability to all open arms available at the site using a centrally-managed computer-generated random sequence. At one site (Bolton) infliximab was unavailable as an intervention. Randomisation was stratified by minimisation with location of patient at the time of randomisation, ward versus ICU, as the sole stratification factor. Although clinical staff were aware of treatment allocation, aggregate outcomes were not provided to them, the trial management committee or the trial steering committee.

### Procedures

Participants assigned to namilumab received a single intravenous (IV) dose of 150mg given over 1 hour on day 1. Those receiving infliximab (Remsima) had a single IV dose of 5 mg/kg over 2 hours on day 1. Participants were followed for 28 days. Blood tests were taken on days 1, 3, 5, 7, 9 and 14 until truncated by discharge or death. Physiological measures were collected until day 14, discharge, or death, and included the ratio of the oxygen saturation to fractional inspired oxygen concentration (SpO_2_/FiO_2;_ SF ratio). The World Health Organisation (WHO) Clinical Progression Improvement Scale was assessed daily for 28 days on a 1-10 scale (online supplementary Table 1) where 1 is asymptomatic, 4 is hospitalised without oxygen, 6 is hospitalised with non-invasive ventilation or high-flow nasal oxygen, 7 is hospitalised with mechanical ventilation and 10 is death; data for level 0 (no viral load detected) was not collected.^20^ If a patient was discharged earlier than day 28 this outcome was collected by means of a diary and scheduled telephone calls.

**Table 1.**
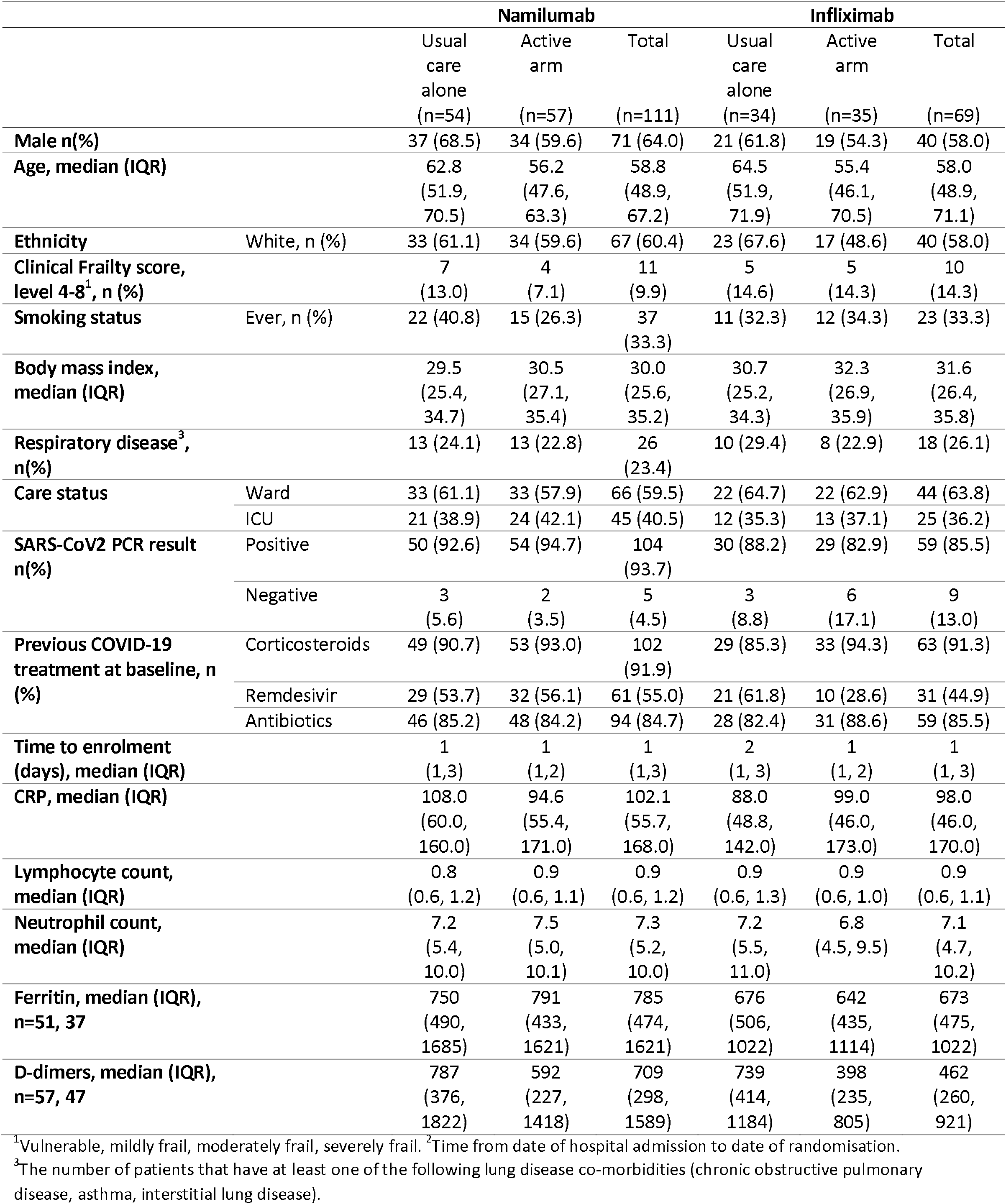
Baseline characteristics

### Outcomes

The primary objective of the trial was to investigate whether candidate treatments could reduce inflammation compared to usual care alone, in order to prioritise drugs to be evaluated in phase 3 trials. The primary outcome measure was CRP, collected over time until day 14. Published data indicate that CRP levels and trajectory are strongly associated with clinical outcomes including respiratory failure and death as well as with lung changes observed on CT.^11^ With the objective of having a rapid, biologically-driven efficacy signal using continuous, readily available data and a small sample size, we had initially chosen the oxygen saturation to fraction of inspired oxygen ratio (SF ratio) as the primary outcome. However, subsequent modelling of data from a large cohort of patients hospitalised in the first wave, indicated that the SF ratio might not be a viable outcome measure of sickness. This led to an early change in primary outcome to CRP, before any analysis of trial data, as previously described.^11^

Secondary outcome measures included the WHO Clinical Progression Scale as a principal clinical efficacy measure as well as hospital survival status and hospital free days, all assessed up to day 28. Hospital free days were defined as the number of days between date of hospital discharge to day 28, with patients who died or who were alive in hospital on day 28 being incorporated as 0 hospital free days. Physiological outcomes measured up to day 14 or discharge, if earlier, included the SF ratio.

Safety data were survival status and adverse events defined by the Common Terminology Criteria for Adverse Events (CTCAE), version 4.03 which fulfilled one of the following criteria: grade ≥ 3, secondary infection or allergic reaction.

### Statistical analysis

The data were analysed according to a pre-specified Statistical Analysis Plan. Each intervention arm was compared against the control group independently, including only control patients for whom that intervention was a randomisation option. For the primary endpoint of CRP, we used Bayesian multi-level regression models that allowed for nesting of the repeated measures data within patient, and non-linear responses, implemented using brms.^21^ Posterior probabilities for the treatment/time interaction covariates were used to conduct decision making at interim analyses, specifically the probability that the covariate was <0 indicating a positive treatment effect in the direction of the intervention as per the model formulation The fitted models incorporated population-level effects for both the intercept and time, random effects for the intercept and time for patient, and fixed effects for age, location (ward/ICU), a main treatment effect, a treatment-time interaction, a treatment-location interaction and a higher order time term.

For the WHO scale, we used Bayesian longitudinal ordinal regression models, implemented using brms,^21^ including in the model formulation fixed effects for location, age, a main treatment effect and a treatment-time interaction and random effects for both the intercepts and time for patient. For consistency with other trials, we also calculated the time to a two-point improvement for this outcome. Results for other outcome measures were not modelled; the results are summarised graphically or tabulated.

Because it is difficult to summarise the results of complex models with a single statistic, we present for the aforementioned models conditional probability plots, which show the predicted values of the natural logarithm of CRP, and, for the WHO scale, the predicted probability of being in each of the WHO outcome categories, conditioned on model parameter values. This enables an easy to interpret visualisation of effect of treatment on these outcomes through time.

Given the lack of any frequentist hypothesis testing there are no reported P-values. Where relevant we include estimates of uncertainty for any point estimates at the stated confidence/probability level typically 95%,

Interim analyses were planned every 20 participants per arm up to 60 participants, and CRP data was considered by the data monitoring committee (DMC) in the context of the emerging safety data to make a recommendation as outlined below:

a. If there is strong evidence of an additional anti-inflammatory effect (CRP) and a satisfactory safety profile consider progression to clinical endpoint evaluation whether in this trial or in another one;
b. If there is no evidence of additional biological effect or an unfavourable safety signal, then terminate arm and do not proceed.

Success was defined as a 90% probability of an intervention arm being better than usual care in reducing CRP as per the posterior probability for the treatment-time interaction covariate outlined above, whereas less than 50% probability defined futility. However, the size of effect and the totality of data were reviewed before recommending adoption by a phase III platform. The operating characteristics, based on a simpler analysis of area under the curve for sequential CRP data, have been previously published,^11^ and indicated a mean sample size of between 43 and 70 patients per comparison.

Pre-planned subgroup analyses were conducted to ascertain the effect of treatment on the primary outcome measure in participants recruited from ward and ICU, and with non-severe and severe disease at baseline, with severe defined as requiring non-invasive or invasive ventilation. The effect of age was also studied.^12^

The primary outcome was analysed on a modified intention to treat population, which included all participants who received trial treatment and had a baseline and at least one post treatment CRP measurement.

The modified intention to treat population for secondary outcomes included all patients who received any trial treatment and with available data for the respective outcome. The safety population included all patients in the usual care arm and all patients who had received a trial intervention in the active arms.

An independent data monitoring committee (DMC) reviewed unblinded data at interim analyses to advise the Trial Steering Committee on whether the trial data (and results from other relevant research), justified the continuing recruitment of further patients. The DMC operated in accordance with a trial-specific charter based on the template created by the Damocles Group. Statistical analysis was conducted in Stata 16 and R Version 4.0.3. The ISRCTN registry number is 40580903.

### Role of the funding source

The funder of the study had no role in study design, data collection, data analysis, data interpretation, or writing of the report. All authors had full access to all the data in the study and had final responsibility for the decision to submit for publication.

## Results

Between 15^th^ June 2020 and 18^th^ February 2021 we assessed 295 patients for eligibility and randomised 146 participants to usual care (n=54), namilumab (n=57) and infliximab (n=35) (Figure 1). Following a DMC review on 21^st^ January 2021 that made recommendations on both arms based on primary outcome analysis, the Trial Steering Committee advised stopping the infliximab arm for futility (probability of benefit 21%) but to continue to recruit to usual care and namilumab, which met criteria for success (probability of benefit 99%), in order to collect further secondary outcome clinical data. A subsequent DMC meeting on 23^rd^ February 2021, advised closing the remaining arms as the trial had reached a natural stopping point and recent changes to standard of care with routine use of tocilizumab would affect conduct of the trial. In total 9 patients withdrew post-randomisation but before treatment and were not included in the analysis: 5 participants at their own or a relative’s request (1 namilumab and 4 infliximab), 1 patient in the usual care group at the request of the treating physician, 1 patient in the infliximab group was reassessed as not having COVID-19, 1 patient in the namilumab group due to initial non-disclosure of information that met an exclusion criterion, and 1 patient in the infliximab group who was withdrawn before treatment when the DMC recommendation to stop the arm was made known.

**Figure 1.**
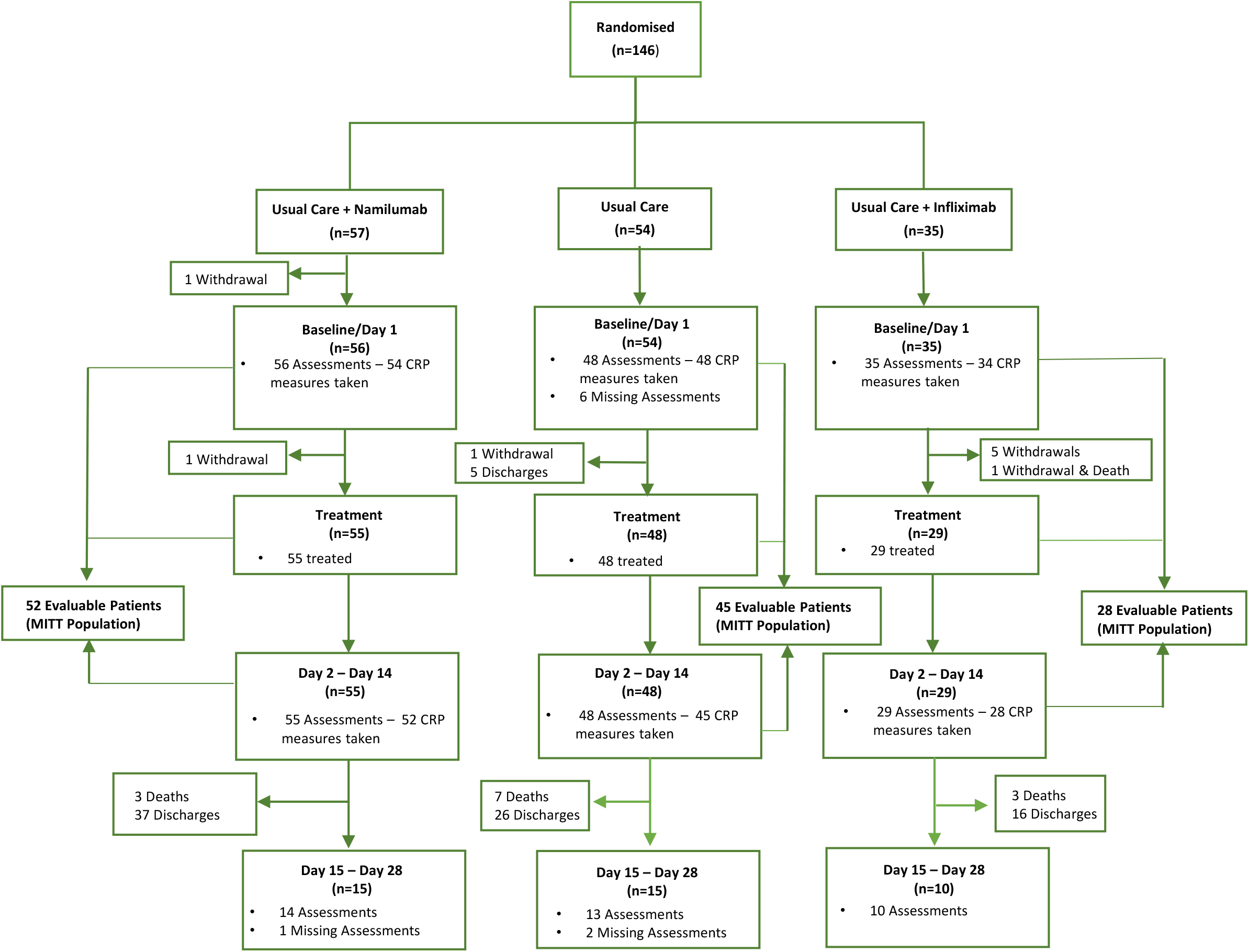
Trial profile indicating number of subjects evaluable for the primary outcome.

Table 1 shows the baseline characteristics for participants. Overall, groups were evenly matched although fewer patients in the infliximab group had remdesivir at enrolment. Most participants had a positive PCR assay for SARS-CoV2. For the usual care and namilumab comparison, 51 (94.4%) and 53 (93.0%) were receiving oxygen, 16 (31.4%) and 19 (35.9%) also receiving high-flow nasal oxygen or CPAP, and 11 (21.6%) and 11 (20.8%) intubated and mechanically ventilated. Almost all patients received dexamethasone as part of usual care at enrolment, and around half received remdesivir. Subsequent to enrolment, all patients bar one in the namilumab group received dexamethasone, 36 (67.9%) and 37 (67.3%) patients in the usual care and namilumab arms received remdesivir, and 8 received tocilizumab (n=3 and n=5 in the usual care and namilumab comparison respectively). For the infliximab comparison, all patients received dexamethasone, 26 (78.8%) and 16 (53.3%) received remdesivir before or following randomisation, and 3 patients received tocilizumab (n=2 and n=1 in usual care and infliximab respectively).

The following patients were evaluable for the primary outcome: 45 and 52 for the usual care alone versus namilumab comparison respectively, and 29 and 28 for the usual care versus infliximab comparison respectively. At the whole population level, and consistent with our previous findings and published data, CRP over time was related to the outcomes of discharge, death and continued hospitalisation at day 28 (supplementary Figure 1). Analysis of the primary outcome showed a 97% probability that namilumab plus usual care was superior to usual care alone in reducing CRP over time (Figure 2). Model fitted values were in good agreement with raw data and indicated a 9% reduction in CRP over usual care alone for each day of follow-up. This effect was consistent in ward and ICU groups based on location at randomisation as visualised in the conditional effects plots (Figure 2), and also in ‘severe’ and ‘non-severe’ patients at baseline (Supplementary Figure 2), where severe was defined as use of non-invasive or invasive ventilation. The effect of namilumab on CRP was independent of age (Supplementary Figure 3). The probability of infliximab being superior to usual care alone was 15%. This lack of effect was consistent across ward and ICU groups (Figure 2) and severe and non-severe disease (supplementary Figure 2). Effects of namilumab and infliximab on CRP were also consistent with an area-under-the curve analysis (data not shown).

**Figure 2.**
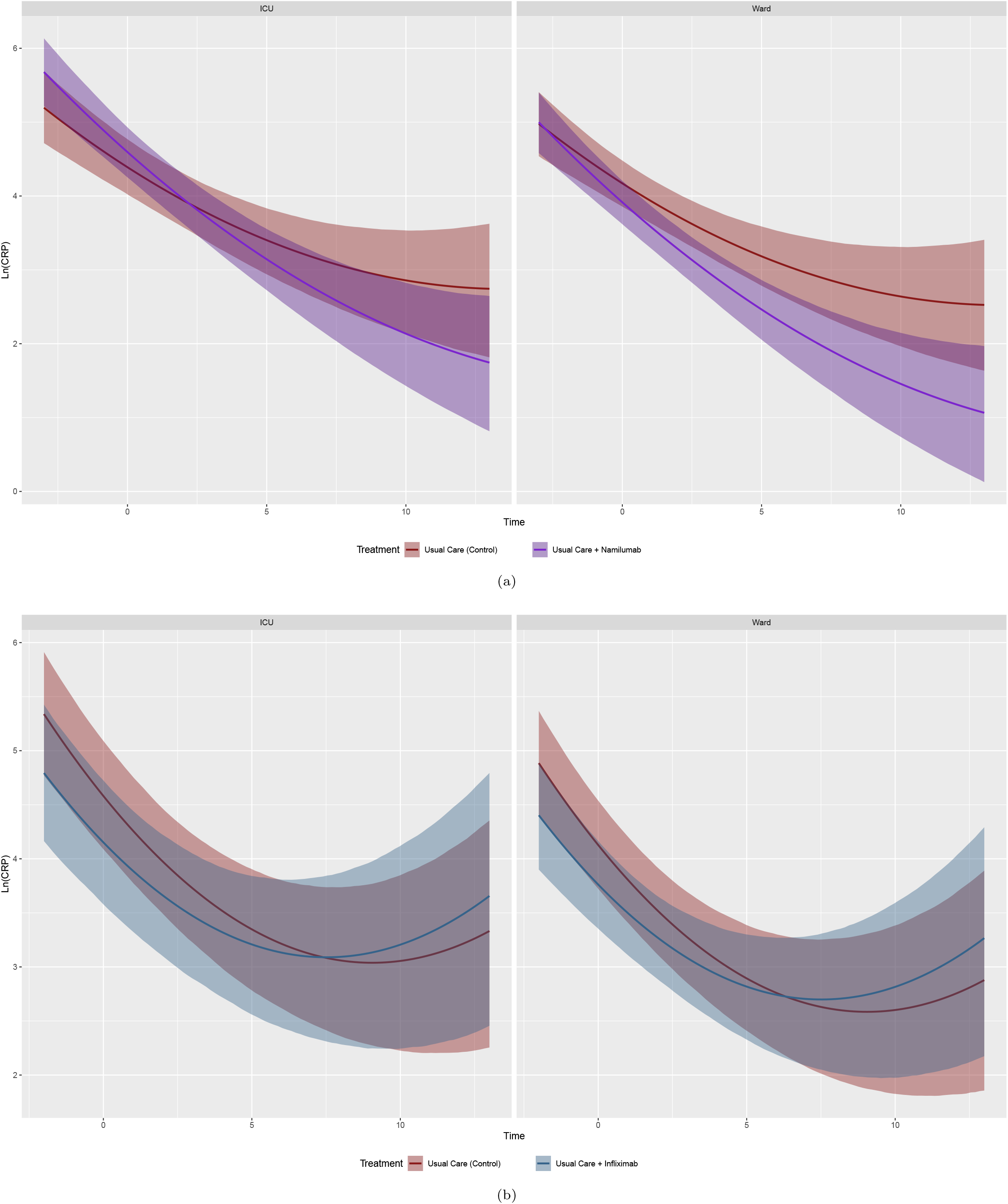
Conditional effects plots of the natural logarithm of CRP modelled over time in days in patients recruited in ward and ICU for namilumab (A) and infliximab (B).

**Figure 3.**
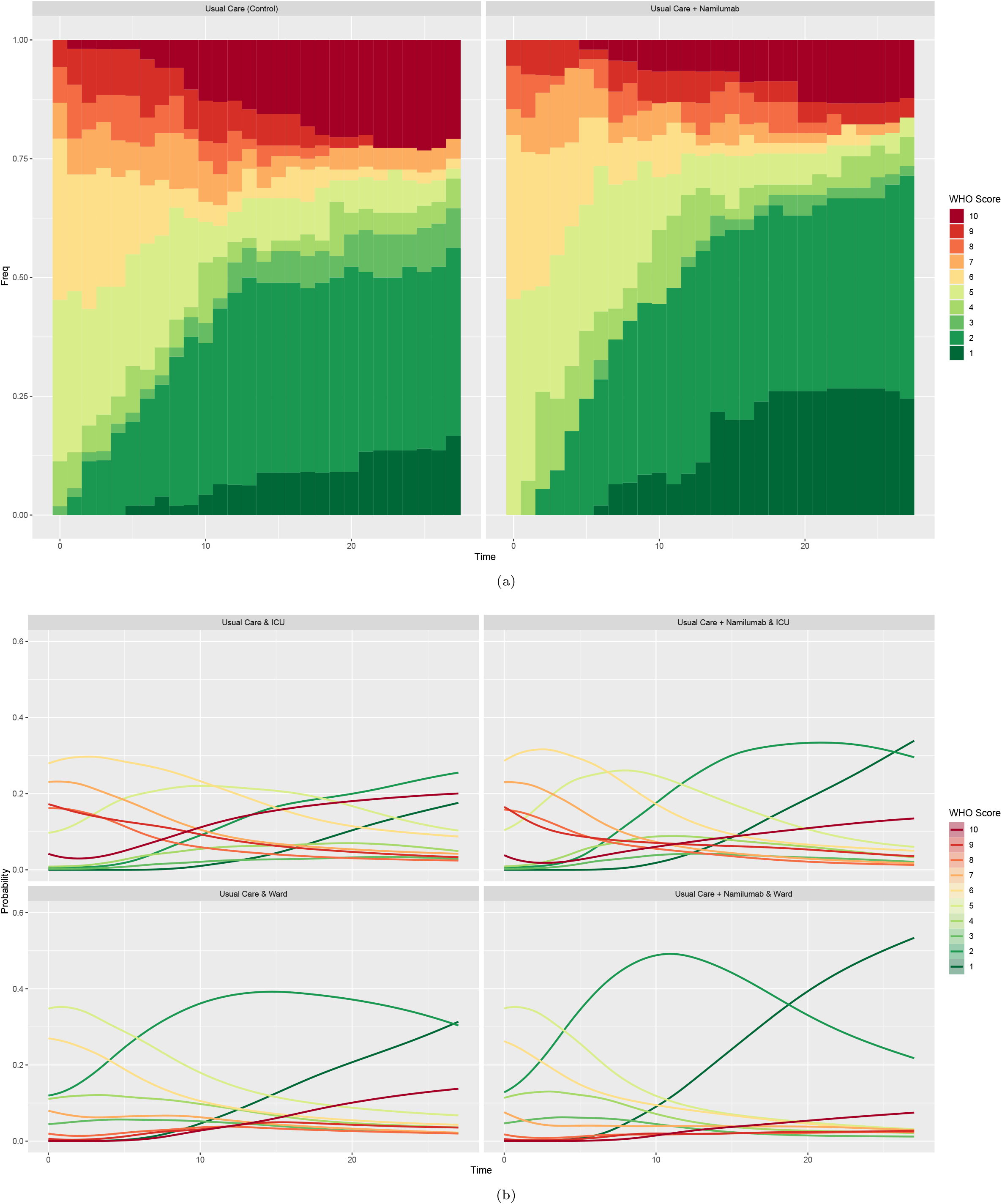
WHO clinical progression score over 28 days for usual care versus namilumab. A, stacked bar chart of raw data for whole population eligible for comparison. B, conditional effects plots of WHO score modelled over time in days showing the probability of being at each level on each day for patients recruited in ICU and ward.

Amongst secondary endpoints, the principal efficacy outcome was the 1-10 point WHO clinical progression scale. For the modified intention-to-treat comparisons between usual care and namilumab, data were available for 53 and 55 patients respectively. Figure 3 shows the proportion of patients at each WHO scale level over 28 days as well as the conditional modelled probabilities of being at each level over time for ward and ICU. In the namilumab arm for patients recruited from both ward and ICU, the probability of having lower scores is consistently increased over time in comparison with usual care. For example, the arms were similar at baseline but by day 28, the probability of discharge (WHO levels 1-3 combined) was 47% and 64% for ICU and ward patients on usual care, versus 66% and 77% for patients treated with namilumab (supplementary Table 2). At day 14, the probability of an ITU patient still needing non-invasive ventilation, invasive ventilation or to have died (WHO ≥ 6) was 54% in the usual care arm vs. 36% in the namilumab arm. Time to two point improvement was also seen to be shorter in the namilumab arm (Table 2 and supplementary Figure 4). Comparable improvements on WHO scale were not observed with infliximab (Figure 4 and supplementary Table 3). The median hospital free days for usual care and namilumab were 17 (IQR 0, 23) and 20 (IQR 3, 23) respectively, and for usual care and infliximab, 17 (0, 23) and 17 (3, 23).

**Table 2.**
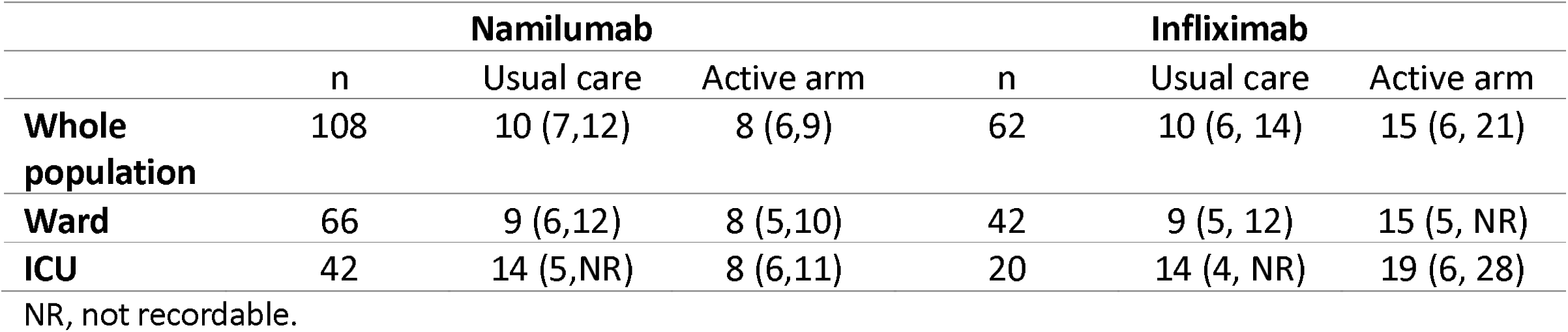
Median time in days (95% CI) to a two point improvement in the WHO clinical progression scale, for overall and subgroups for both drugs. NR, not recordable.

**Table 3.** Hospital discharge status at day 28. Data was available on all patients, n(%).

|  | Status | Namilumab |  |  | Infliximab |  |  |
| --- | --- | --- | --- | --- | --- | --- | --- |
|  |  | Usual care<br>(n=54) | Active arm<br>(n=55) | Overall<br>(n=109) | Usual care<br>(n=34) | Active arm<br>(n=29) | Overall<br>(n=63) |
| **Whole population** | Discharge | 33<br>(61.1) | 43<br>(78.2) | 76<br>(69.7) | 22<br>(64.7) | 22<br>(75.9) | 44<br>(69.8) |
|  | In hospital | 11<br>(20.4) | 6<br>(10.9) | 17<br>(15.6) | 7<br>(20.6) | 3<br>(10.3) | 10<br>(15.9) |
|  | Death | 10<br>(18.5) | 6<br>(10.9) | 16<br>(14.7) | 5<br>(14.7) | 4<br>(13.8) | 9<br>(14.3) |
| **Ward** | Discharge | 28<br>(84.8) | 29<br>(87.9) | 57<br>(86.4) | 19<br>(86.4) | 16<br>(80.0) | 35<br>(83.3) |
|  | In hospital | 4<br>(12.1) | 2<br>(6.1) | 6<br>(9.1) | 2<br>(9.1) | 1<br>(5.0) | 3<br>(7.1) |
|  | Death | 1<br>(3.0) | 2<br>(6.1) | 3<br>(4.5) | 1<br>(4.5) | 3<br>(15.0) | 4<br>(9.5) |
| **ICU** | Discharge | 5<br>(23.8) | 14<br>(63.6) | 19<br>(44.2) | 3<br>(25.0) | 6<br>(66.7) | 9<br>(42.9) |
|  | In hospital | 7<br>(33.3) | 4<br>(18.2) | 11<br>(25.6) | 5<br>(41.7) | 2<br>(22.2) | 7<br>(33.3) |
|  | Death | 9<br>(42.9) | 4<br>(18.2) | 13<br>(30.2) | 4<br>(33.0) | 1<br>(11.1) | 5<br>(23.8) |

**Figure 4.**
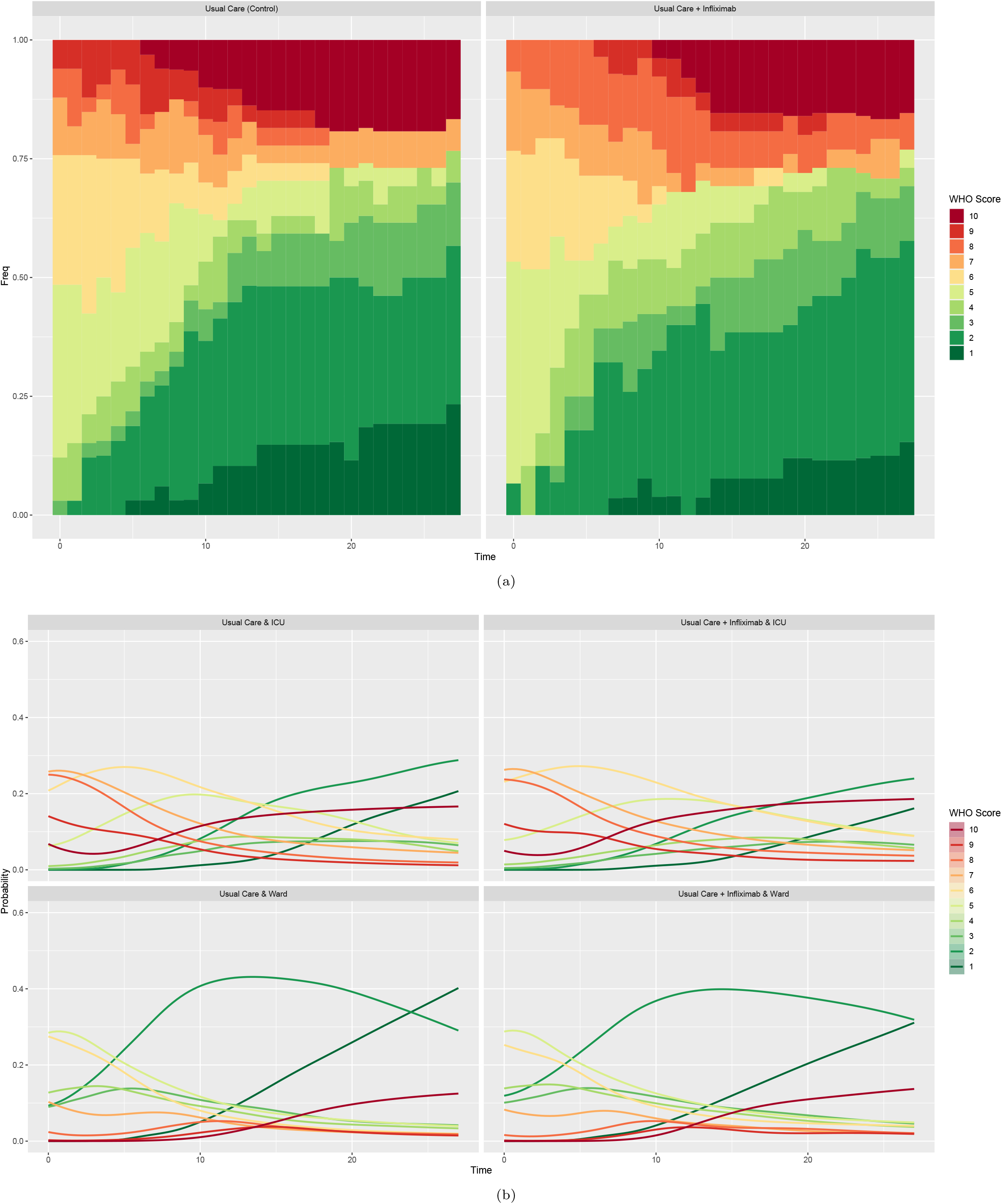
WHO clinical progression score over 28 days for usual care versus infliximab. A, stacked bar chart of raw data for whole population eligible for comparison. B, conditional effects plots of WHO score modelled over time in days showing the probability of being at each level on each day for patients recruited in ICU and ward.

By day 28, there were fewer deaths and more discharges in the namilumab group with 43 (78.2%) participants discharged, 6 (10.9%) still in hospital and 6 (10.9%) dead, compared to 33 (61.6%), 11 (20.4%), and 10 (18.5%) for usual care alone (Table 3). Interestingly, despite the challenges we described in modelling the SF ratio, trends to improvement in oxygenation status were observed with namilumab (supplementary Figure 5).

For the namilumab and usual care comparison, a total of 279 adverse events were reported in 59 of the 109 patients in the safety population (54.1%; 134 events in n=30 and 145 events in n=29 for namilumab and usual care respectively). Of these, 131 (90.3%) and 103 events (76.9%) events were grade 3 or above for usual care and namilumab respectively. Infections were more common in the namilumab group (20 events) compared with usual care (10 events). Table 4 shows adverse events that were grade ≥ 3, secondary infection or allergic reaction, for which more than one event occurred. There were 10 serious adverse events in each of the usual care and namilumab groups respectively. All except one of the namilumab SAEs were considered unrelated, the related case being a re-admission with bacterial pneumonia 26 days after receiving namilumab and on a background of a prolonged admission for social reasons and known COPD.

**Table 4.** Adverse events that are CTCAE grade ≥3, secondary infection or allergic reaction. Only events occurring at least twice within an active drug/usual care comparison are shown. Data shown are number of adverse event occurrences (number of patients affected).

|  | **Namilumab** |  | **Infliximab** |  |
| --- | --- | --- | --- | --- |
|  | Usual care | Active arm | Usual care | Active arm |
| **Anaemia** | 10 (6) | 10(6) | 8 (5) | 2 (2) |
| **Sinus bradycardia** | 2 (1) | 0 (0) | 2 (1) | 0 (0) |
| **Multiorgan failure** | 3 (3) | 1 (1) | - | - |
| **Covid pneumonia/pneumonitis** | 5(5) | 4(4) | 2 (2) | 2 (2) |
| **Lung infection** | 2(2) | 1(1) | - | - |
| **Pleural infection** | 2(1) | 0(0) | 2 (1) | 0 (0) |
| **Sepsis** | 1(1) | 2(2) | - | - |
| **Raised ALT** | 3(3) | 5(5) | 1 (1) | 1 (1) |
| **Raised Troponin I** | 0(0) | 2(1) | - | - |
| **Raised Creatinine** | 2(2) | 4(3) | 2(2) | 2(1) |
| **Raised CRP** | 5(4) | 2(2) | 5(4) | 0(0) |
| **Raised d-dimers** | 6(5) | 3(3) | 5(4) | 1(1) |
| **Raised ferritin** | 7(6) | 5(5) | 5(4) | 11(7) |
| **Low lymphocytes** | 16(12) | 5(3) | 11 (8) | 4 (2) |
| **Raised monocytes** | 0(0) | 3(2) | - | - |
| **Raised neutrophils** | 9(5) | 5(3) | 9 (5) | 4 (4) |
| **Raised white cells** | 9(6) | 7(4) | 9 (6) | 9 (5) |
| **Low platelets** | 1(1) | 1(1) | - | - |
| **Raised urea** | 9(7) | 11(5) | 8(6) | 8(5) |
| **Raised potassium** | 6(5) | 1(1) | 4(3) | 2(1) |
| **Raised sodium** | 1(1) | 2(1) | - | - |
| **Raised triglycerides** | 3(3) | 1(1) | 2(2) | 6(4) |
| **Low albumin** | 15 (13) | 11 (7) | 12 (10) | 13 (8) |
| **Low sodium** | 2(2) | 1(1) | 2(2) | 2(2) |
| **ARDS** | - | - | 1(1) | 1(1) |
| **Hypotension** | - | - | 0(0) | 2(2) |

For the infliximab and usual care comparison, a total of 214 adverse events were reported in 37 of the 63 patients in the safety population (59.7%; 112 events in 17 usual care patients and 102 events in 20 infliximab patients). Of these, 101 (90.2%) and 78 (76.5%) were grade 3 or above for usual care and infliximab respectively. There were 7 infection events in usual care and 4 with infliximab. There were 5 serious adverse events in the usual care group and 6 with infliximab, all considered unrelated.

## Discussion

Our trial clearly demonstrated that the addition of namilumab, but not infliximab to usual care, reduced inflammation as measured by CRP in hospitalised patients with COVID-19, when compared to usual care alone. Importantly, the secondary clinical outcomes are consistent and shared the same directionality as the primary outcome for both interventions, despite not being formally powered to assess for such differences. Our proof-of-concept findings with GM-CSF inhibition is consistent with our hypothesis that recruitment and activation of IMM are important in the pathogenesis of severe COVID-19. This is also consistent with published findings from small non-randomised trials,^22,23^ and recent, large randomised trials of other GM-CSF inhibitors in COVID-19. Otilimab showed benefit for the primary endpoint of being alive and free of respiratory failure at day 28 in a predefined subgroup of patients aged 70 or over.^24^ Lenzilumab, given as a three dose course in non-ventilated hospitalised patients, showed benefit over standard care in the primary outcome of survival without ventilation, an effect that seemed more pronounced in patients aged 85 or under and with CRP <150 mg/L.^25^ Our data suggest the effect of a single dose of namilumab on CRP and WHO score is independent of age, although this requires confirmation in larger studies.

In the absence of large treatment effects, small trials using traditional clinical outcomes may give inconclusive or contrary findings in COVID-19, as exemplified by earlier studies of tocilizumab. The CATALYST trial was designed to use a repeatedly collected continuous measure of CRP with a Bayesian adaptive approach that we predicted would require a smaller sample size to show evidence of efficacy or futility. CRP levels, including the rate of decline, have been associated with clinical outcome in COVID-19 (reviewed in^11^) and we hypothesised that an immunomodulatory agent unable to alter CRP would be a less promising candidate to take forward into phase 3 trials. In the face of many options for repurposing immunomodulatory therapies in COVID-19, we contend that such a prioritisation approach will make the most efficient use of phase 3 resource and accelerate development of effective drugs.

In contrast to the observed effect of namilumab, we could not demonstrate a comparable benefit on CRP with infliximab and the arm was stopped for futility. TNF is in important pro-inflammatory cytokine produced by macrophages as well as other cell types, with context-dependent pleiotropic effects including further activation of IMM and up-regulation of inflammatory mediators such as IL-6. One previous non-randomised study of infliximab suggested potential efficacy, albeit with significant limitations including small sample size, use of historical controls, and being conducted prior to routine use of corticosteroids.^26^ This, together with circumstantial data, justified our inclusion of infliximab.^18^ However, although TNF inhibitors are widely used in inflammatory diseases, not all IMID are responsive, and TNF itself may suppress certain pro-inflammatory factors that may be relevant to COVID-19 such as type 1 interferon expression and Th17 cell differentiation.^27^ Inhibition of such cross-regulatory effects may underlie our negative findings, or simply indicate that TNF is not on a critical path to driving inflammatory responses as measured by CRP in patients hospitalised with COVID-19. GM-CSF inhibition might also have an additional benefit in retarding neutrophil recruitment and activation that may be of importance in the pathogenesis of severe COVID-19 and acute respiratory distress syndrome.^28^ Our safety data suggest that the lack of response to infliximab is not due to an increase in secondary infections. Whilst we cannot exclude the possibility of benefit being seen in a subset of patients or in larger studies, the clear divergence in primary outcome is broadly reflected in the secondary clinical findings and justifies the prioritisation of GM-CSF inhibition over TNF inhibition for further study in hospitalised COVID-19 patients.

GM-CSF has an important role in the differentiation of alveolar macrophages, and consequently in surfactant clearance, as well as being an important survival factor for lung epithelial cells. Absence of GM-CSF signalling, through genetic defect in the receptor or very high levels of polyclonal autoantibodies to GM-CSF, have been associated with pulmonary alveolar proteinosis (PAP). PAP has been an adverse event of special interest in previous clinical trials of GM-CSF inhibitors but, to our knowledge, has never been observed. It is important to note, (i) that therapeutic monoclonal antibodies will not completely inhibit GM-CSF signalling which appears to be a requirement for PAP,^29^ but rather will down-regulate excessive pathway activation, (ii) lack of GM-CSF does not prevent macrophage uptake of surfactant as much as its catabolism, therefore the effect of short-term inhibition is likely to be less pronounced on surfactant clearance when compared with long-term inhibition, (iii) down regulation of monocyte activation, which is the aim of GM-CSF inhibition, should itself lead to a reduction in alveolar epithelial cell damage in COVID-19. However it is also important to note an opposing view that administration of GM-CSF might have therapeutic benefits and the results of clinical trials of inhaled and intravenous sargramostim are awaited.^30^

Our study has a number of limitations. Similar to many other trials in COVID-19 we did not use a placebo control. However, the discordant results of the two active arms, when compared to usual care, as well as the objective nature of CRP data, suggest this does not explain the positive findings we observed with namilumab. Our sample size is too small for a definitive assessment of clinical outcomes and further studies are required for this as well as to understand better the population that may benefit most. Our results may not generalise to hospitalised patients without evidence of pneumonia or raised CRP or patients not requiring hospitalisation. Our data emphasise the need to monitor secondary infections in future COVID-19 trials, particularly given the use of combination immune-modulating treatments.

Despite the advances of dexamethasone and tocilizumab in COVID-19, mortality amongst patients with severe disease remain high.^2^ There therefore remains considerable unmet medical need, and data pointing to the role of both IMM and GM-CSF in severe COVID-19, together with our findings reported here, strongly suggest that targeted GM-CSF inhibitors such as namilumab should be further investigated in hospitalised patients with COVID-19.

**Research in Context**

**Evidence before this study**

We searched Pubmed and medRxiv on 10^th^ May 2021, using the following search terms [(randomised OR trial) AND (anti-GM-CSF OR namilumab OR mavrilimumab OR otilimab OR lenzilumab OR gimsilumab OR TJ003234 OR anti-TNF OR infliximab OR adalimumab OR etanercept OR golimumab OR certolizumab) AND (COVID* OR SARS-CoV-2 OR SARS-CoV)]. Two small non-randomised studies with drugs targeting GM-CSF or its receptor (lenzilumab and mavrilimumab) and one study with a TNF inhibitor (infliximab) have all suggested potential efficacy but with significant limitations of small sample size, use of historical controls, and being conducted prior to routine use of corticosteroids. One RCT with mavrilimumab was small and inconclusive. Two larger RCTs with other anti-GM-CSF inhibitors have recently been published. Otilimab showed benefit for the primary endpoint of being alive and free of respiratory failure at day 28 in a predefined subgroup of patients aged 70 or over. Lenzilumab, given as a three dose course, in non-ventilated hospitalised patients showed benefit over standard care in the primary outcome of survival without ventilation, an effect that seemed more pronounced in patients aged 85 or under and with CRP <150 mg/L. We identified no published randomised trials of TNF inhibitors in COVID-19.

**Added value of this study**

This is the first randomised trial of namilumab and infliximab in COVID-19. We found that both drugs were safe and that namilumab, but not infliximab, showed proof of concept evidence of reduction in inflammation as measured by CRP in hospitalised patients with COVID-19 pneumonia. Secondary clinical outcomes were concordant with the primary outcome, with trends to improvement in patients recruited from both ward and ICU.

**Implications of all the available evidence**

Consistent with emerging evidence implicating GM-CSF and inflammatory monocytes/macrophages in the pathogenesis of severe COVID-19, namilumab improved both biological and clinical outcomes. It should be prioritised for further study in COVID-19.

## Data Availability

Participant data and the associated supporting documentation will be available within six months after the publication of this manuscript. Details of our data request process are available on the Cancer Research UK Clinical Trials Unit (CRCTU) website. Only scientifically sound proposals from appropriately qualified research groups will be considered for data sharing. The decision to release data will be made by the CRCTU Directors Committee who will consider the scientific validity of the request and the qualifications and resources of the research group and the views of the Chief Investigator and the Trial Steering Committee, consent arrangements and the practicality of anonymising the requested data and contractual obligations. A data sharing agreement will cover the terms and conditions of the release of trial data and will include publication requirements and authorship and acknowledgements and obligations for the responsible use of data. An anonymised encrypted dataset will be transferred directly using a secure method and in accordance with the University of Birmingham IT guidance on encryption of datasets.

## Contributors

BAF, TV, MR, TW, DP, AR, RS. DRT, JB, SG, DR and PK conceived the study. BAF, TV, DS, MR, TW, DP, AR, RS, DRT, JB, HM, LH, PNN, SG, DR and PK designed the clinical trial. BAF and DP were arm leads for namilumab and MR and DR were arm leads for infliximab. TV, TW, JS, DP, MSB, GC, NM, ZG, MPW, JP and AR recruited patients and/or collected data. DS, CG and SG conducted the statistical analysis. BAF drafted the manuscript which all authors revised and approved for submission.

## Declaration of interests

BAF has undertaken consultancy for Novartis, BMS, Servier, Galapagos and Janssen and received research funding from Servier and Galapagos; MR is currently undertaking a Senior Clinical Fellowship financed by Roche; PK has undertaken consultancy for BMS, and AstraZeneca, and has received research funding from Bayer and Pfizer; DR is a former employee of GSK; all are unrelated to this trial. All other authors declare no competing interests.

## Data sharing

Participant data and the associated supporting documentation will be available within six months after the publication of this manuscript. Details of our data request process are available on the Cancer Research UK Clinical Trials Unit (CRCTU) website. Only scientifically sound proposals from appropriately qualified research groups will be considered for data sharing. The decision to release data will be made by the CRCTU Director’s Committee, who will consider the scientific validity of the request, the qualifications and resources of the research group, the views of the Chief Investigator and the Trial Steering Committee, consent arrangements, the practicality of anonymising the requested data and contractual obligations. A data sharing agreement will cover the terms and conditions of the release of trial data and will include publication requirements, authorship and acknowledgements and obligations for the responsible use of data. An anonymised encrypted dataset will be transferred directly using a secure method and in accordance with the University of Birmingham’s IT guidance on encryption of datasets.

## Acknowledgements

This trial is supported by the Medical Research Council (MRC) grant number MC_PC_20007. SG is supported by a Senior Investigator Award from the National Institute of Health Research. Staff at the CRCTU are supported by core funding grants from Cancer Research UK (C22436/A25354), the NIHR Biomedical Research Centre (BRC-1215-20009), The Kennedy Trust for Rheumatology Research as part of the Arthritis – Trials Acceleration Programme (KENN161704), and Innovate UK as part of the Midlands – Wales Advanced Therapy Treatment Centres (104232). This paper presents independent research supported by the NIHR Birmingham Biomedical Research Centres at Birmingham, Oxford, Imperial College London and University College London. GC is supported by a NIHR Research Professorship. The views expressed are those of the authors and not necessarily those of the NHS, the NIHR or the Department of Health and Social Care. Namilumab was provided free of charge by Izana Bioscience, Oxford, UK (now part of Roivant). Infliximab is being provided free of charge by Celltrion.

## Notes

### Clinical Trial

ISRCTN 40580903

### Author Declarations

East Midlands-Nottingham 2 Research Ethics Committee (20/EM/0115)

