## Supplementary material for "Namilumab or infliximab compared to standard of care in hospitalised patients with COVID-19 (CATALYST): a phase 2 randomised adaptive trial": Supplemenary Data

**Additional CATALYST Investigators**

**Data Monitoring Committee**

| Member | DMC Role | Contact Information |
|---|---|---|
| Professor Adam Hill | Chair | University of Edinburgh |
| Professor Christina Yap | Independent Statistician | The Institute of Cancer Reserarch, London |
| Professor Anthony Gordon | | Imperial College London |

**Trial Steering Committee**

| Member | TSC Role | Contact Information |
|---|---|---|
| Professor Michael Matthay | Chair | University of California San Francisco |
| Professor Danny McAuley | Deputy Chair | Queen’s University Belfast |
| Mr Andrew Hall | Independent Statistician | University of Leeds |
| Professor Paul Dark | | Manchester University |
| Professor Sir Andrew McMichael | | University of Oxford |

**Scientific Advisory Board**

| Member | | Contact Information |
|---|---|---|
| Professor Philip Newsome | Chair | University of Birmingham |
| Professor Pamela Kearns | | University of Birmingham |
| Professor Tonny Veenith | | University of Birmingham |
| Dr Benjamin Fisher | | University of Birmingham |
| Dr Rowena Sharpe | | University of Birmingham |
| Professor Julian Bion | | University of Birmingham |
| Professor Bryan Williams | | University College London |
| Professor Duncan Richards | | University of Oxford |
| Professor Graham Cooke | | Imperial College London |
| Professor Helen McShane | | University of Oxford |
| Professor Ling-Pei Ho | | University of Oxford |
| Dr Rebecca Turner | | University College London |
| Professor Simon Gates | | University of Birmingham |
| Professor Vincenzo Libri | | University College London |

**Supplementary Table 1 World Health Organisation Clinical Progression Scale**

| Patient State | Descriptor | Score |
|---|---|---|
| Uninfected | Uninfected; no viral RNA detected | 0 |
| Ambulatory | Asymptomatic; viral RNA detected | 1 |
| | Symptomatic; independent | 2 |
| | Symptomatic; assistance needed | 3 |
| Hospitalised; mild disease | Hospitalised; no oxygen therapy | 4 |
| | Hospitalised; oxygen by mask or nasal prongs | 5 |
| Hospitalised; severe disease | Hospitalised; oxygen by NIV or high flow | 6 |
| | Intubated and mechanical ventilation, $pO_2/FiO_2 \geq 150$ or $SpO_2/FiO_2 \geq 200$ | 7 |
| | Mechanical ventilation $pO_2/FiO_2 < 150$ ($SpO_2/FiO_2 < 200$) **or** vasopressors | 8 |
| | Mechanical ventilation $pO_2/FiO_2 < 150$ ($SpO_2/FiO_2 < 200$) **and** vasopressors, dialysis or ECMO | 9 |
| Death | Dead | 10 |

Adapted from WHO Working Group on the Clinical Characterisation and Management of COVID-19 infection. A minimal common outcome measure set for COVID-19 clinical research. Lancet Infect Dis. 2020;20:e192-e197.

**Footnotes for use in CATALYST**

1. If $pO_2$ not available then use the $SpO_2/FiO_2$ ratio instead

2. For $pO_2$ measurements in kPa, use an online calculator e.g. https://www.msdmanuals.com/en-gb/medical-calculators/PaO2_FiO2Ratio.htm to calculate a $pO_2/FiO_2$ ratio equivalent to that obtained with $pO_2$ measured in mmHg, or else consider an equivalent ratio to 200, when dividing $pO_2$ in kPa by $FiO_2$, is 26.7, and an equivalent to 150 is 20.

3. If medically fit for discharge, record status as for ambulatory patient

4. Asymptomatic implies a return to baseline symptomatic state, i.e. no fever, and no cough, shortness of breath, confusion, myalgia, diarrhoea, fatigue, or weakness above what the participant would have experienced on a daily basis before their COVID-19 episode

5. Symptomatic but independent, implies that the participant has some of the additional symptoms as above, but needs no additional help with activities of daily living above what they required prior to their COVID-19 episode.

6. Symptomatic but needs assistance, implies that in addition to having symptoms as above, they require help with activities of daily living i.e. bathing/showering, personal hygiene and combing of hair, dressing, toileting, mobility/transferring and self-feeding, above what they required on a daily basis prior to their COVID-19 episode.

7. Score 0 (uninfected: no viral RNA detected) is not being assessed as part of CATALYST.

**Supplementary table 2**. Point estimates of the probability of being at each level of the WHO Clinical progression score on days 1, 14 and 28 for ward and ICU patients allocated to usual care alone or namilumab plus usual care. Conditional effects data derived from Bayesian longitudinal proportional odds ordinal regression.

| | | | WHO score level | | | | | | | | | |
|---|---|---|---|---|---|---|---|---|---|---|---|---|
| | CareStatus | Day | 1 | 2 | 3 | 4 | 5 | 6 | 7 | 8 | 9 | 10 |
| Usual Care (Control) | Intensive Care Unit | 1 | 0.00 | 0.01 | 0.00 | 0.01 | 0.10 | 0.28 | 0.23 | 0.16 | 0.17 | 0.04 |
| | Intensive Care Unit | 14 | 0.03 | 0.13 | 0.03 | 0.06 | 0.22 | 0.20 | 0.08 | 0.05 | 0.07 | 0.14 |
| | Intensive Care Unit | 28 | 0.18 | 0.26 | 0.03 | 0.05 | 0.10 | 0.09 | 0.04 | 0.02 | 0.03 | 0.19 |
| | On Ward | 1 | 0.00 | 0.12 | 0.04 | 0.11 | 0.35 | 0.27 | 0.08 | 0.02 | 0.01 | 0.00 |
| | On Ward | 14 | 0.09 | 0.39 | 0.05 | 0.08 | 0.14 | 0.08 | 0.05 | 0.04 | 0.04 | 0.04 |
| | On Ward | 28 | 0.31 | 0.31 | 0.02 | 0.04 | 0.07 | 0.04 | 0.02 | 0.02 | 0.03 | 0.14 |
| Usual Care + Namilumab | Intensive Care Unit | 1 | 0.00 | 0.01 | 0.00 | 0.01 | 0.10 | 0.29 | 0.23 | 0.16 | 0.16 | 0.04 |
| | Intensive Care Unit | 14 | 0.06 | 0.26 | 0.04 | 0.09 | 0.21 | 0.12 | 0.05 | 0.04 | 0.07 | 0.08 |
| | Intensive Care Unit | 28 | 0.34 | 0.30 | 0.02 | 0.03 | 0.06 | 0.05 | 0.02 | 0.01 | 0.03 | 0.13 |
| | On Ward | 1 | 0.00 | 0.12 | 0.05 | 0.12 | 0.35 | 0.26 | 0.07 | 0.02 | 0.01 | 0.00 |
| | On Ward | 14 | 0.18 | 0.48 | 0.03 | 0.05 | 0.08 | 0.07 | 0.04 | 0.02 | 0.02 | 0.03 |
| | On Ward | 28 | 0.54 | 0.22 | 0.01 | 0.02 | 0.03 | 0.03 | 0.03 | 0.02 | 0.03 | 0.07 |

**Supplementary table 3**. Point estimates of the probability of being at each level of the WHO Clinical progression score on days 1, 14 and 28 for ward and ICU patients allocated to usual care alone or infliximab plus usual care. Conditional effects data derived from Bayesian longitudinal proportional odds ordinal regression.

| | | | WHO score level | | | | | | | | | |
|---|---|---|---|---|---|---|---|---|---|---|---|---|
| | Care Status | Day | 1 | 2 | 3 | 4 | 5 | 6 | 7 | 8 | 9 | 10 |
| Usual Care (Control) | Intensive Care Unit | 1 | 0.00 | 0.00 | 0.00 | 0.01 | 0.06 | 0.21 | 0.26 | 0.25 | 0.14 | 0.07 |
| | Intensive Care Unit | 14 | 0.03 | 0.15 | 0.07 | 0.08 | 0.18 | 0.18 | 0.09 | 0.05 | 0.04 | 0.14 |
| | Intensive Care Unit | 28 | 0.21 | 0.28 | 0.06 | 0.05 | 0.07 | 0.08 | 0.04 | 0.02 | 0.01 | 0.17 |
| | On Ward | 1 | 0.00 | 0.09 | 0.09 | 0.13 | 0.28 | 0.27 | 0.10 | 0.02 | 0.00 | 0.00 |
| | On Ward | 14 | 0.11 | 0.42 | 0.09 | 0.07 | 0.09 | 0.05 | 0.04 | 0.05 | 0.04 | 0.03 |
| | On Ward | 28 | 0.40 | 0.28 | 0.04 | 0.03 | 0.04 | 0.02 | 0.02 | 0.02 | 0.01 | 0.13 |
| Usual Care + Infliximab | Intensive Care Unit | 1 | 0.00 | 0.00 | 0.00 | 0.01 | 0.08 | 0.23 | 0.26 | 0.24 | 0.12 | 0.05 |
| | Intensive Care Unit | 14 | 0.02 | 0.12 | 0.05 | 0.07 | 0.18 | 0.19 | 0.10 | 0.07 | 0.04 | 0.15 |
| | Intensive Care Unit | 28 | 0.16 | 0.24 | 0.06 | 0.06 | 0.09 | 0.09 | 0.05 | 0.04 | 0.02 | 0.19 |
| | On Ward | 1 | 0.00 | 0.12 | 0.10 | 0.14 | 0.29 | 0.25 | 0.08 | 0.02 | 0.00 | 0.00 |
| | On Ward | 14 | 0.09 | 0.39 | 0.10 | 0.08 | 0.10 | 0.07 | 0.05 | 0.04 | 0.04 | 0.05 |
| | On Ward | 28 | 0.31 | 0.31 | 0.05 | 0.04 | 0.05 | 0.04 | 0.02 | 0.02 | 0.02 | 0.14 |

Figure 1: Supplementary Figure 1. CRP over time in relation to day 28 outcomes of death, discharge, and ongoing hospitalisation within the whole CATALYST modified intention to treat population.

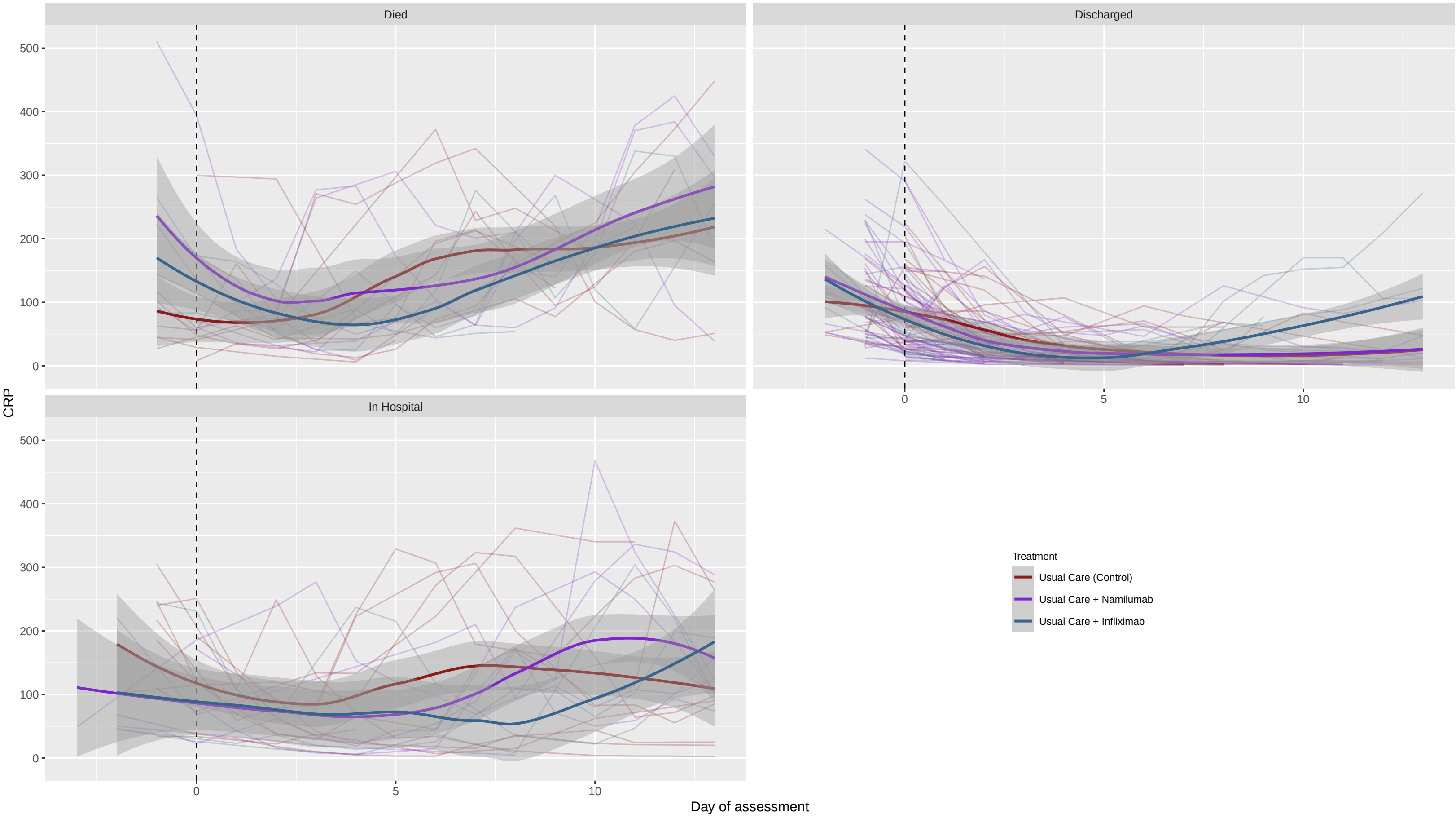

Figure 2: Supplementary Figure 2. Conditional effects plots of CRP modelled over time in patients recruited in with non-severe and severe disease at baseline for namilumab (A) and infliximab (B). Severe disease was defined as the use of non-invasive or invasive ventilation or high flow nasal oxygen at baseline. Time 0 is day 1 of assessment.

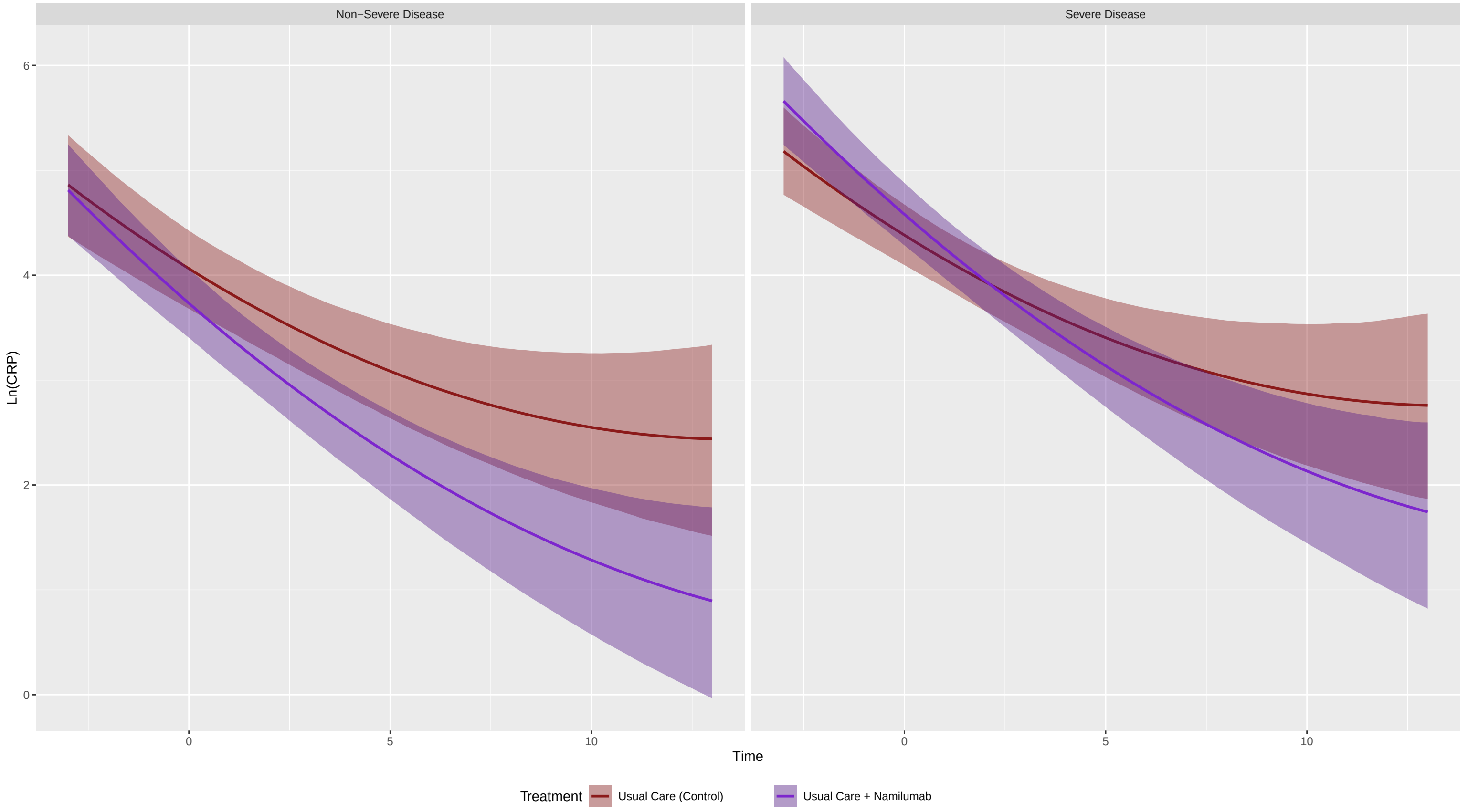


(a)

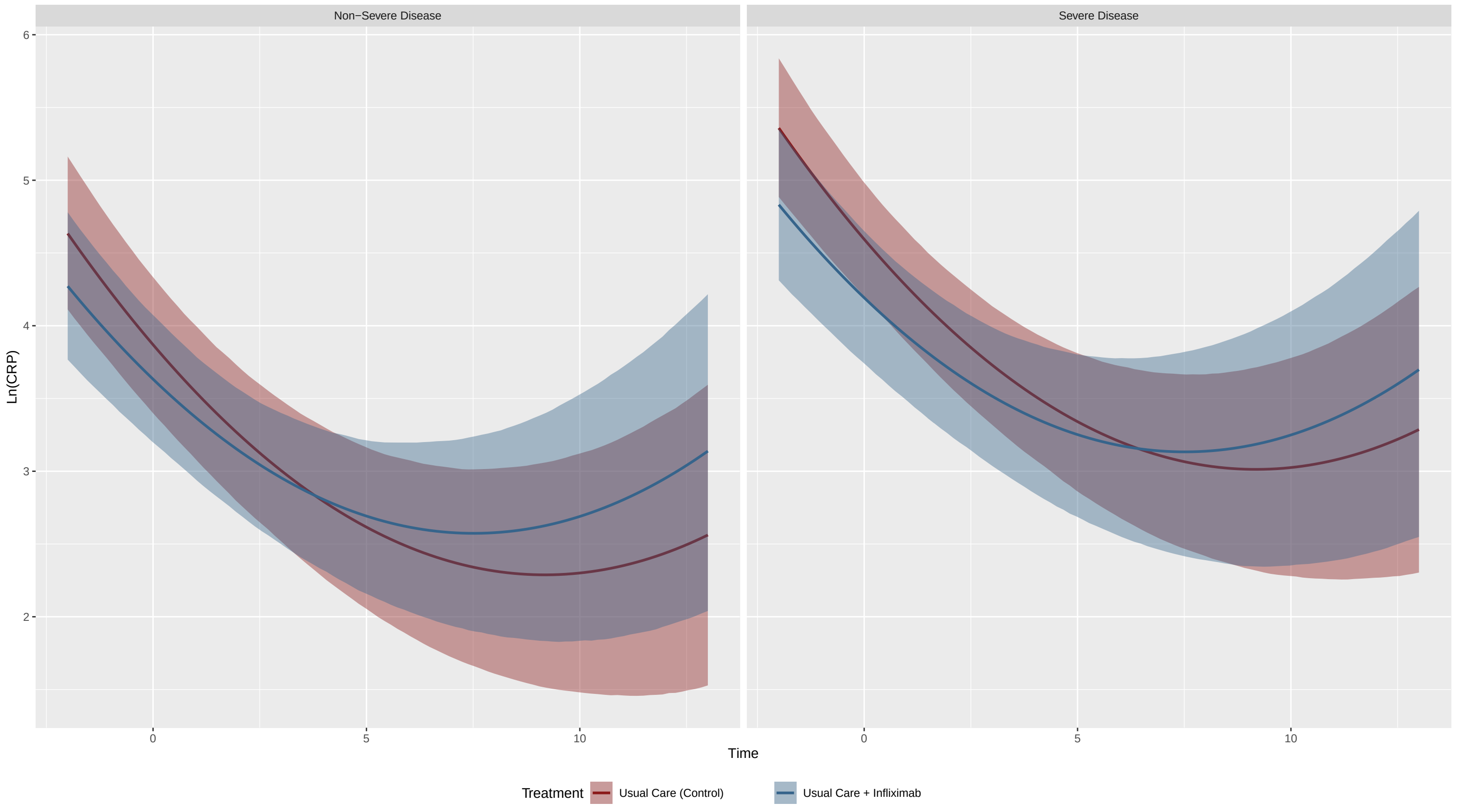


(b)

Figure 3: Supplementary Figure 3. Conditional effects model of CRP in relation to age and treatment at days 0, 7 and 14 in ward and ICU groups. CRP is associated with age but the effect of (A )namilumab (A) and (B) infliximab on CRP is independent of age.

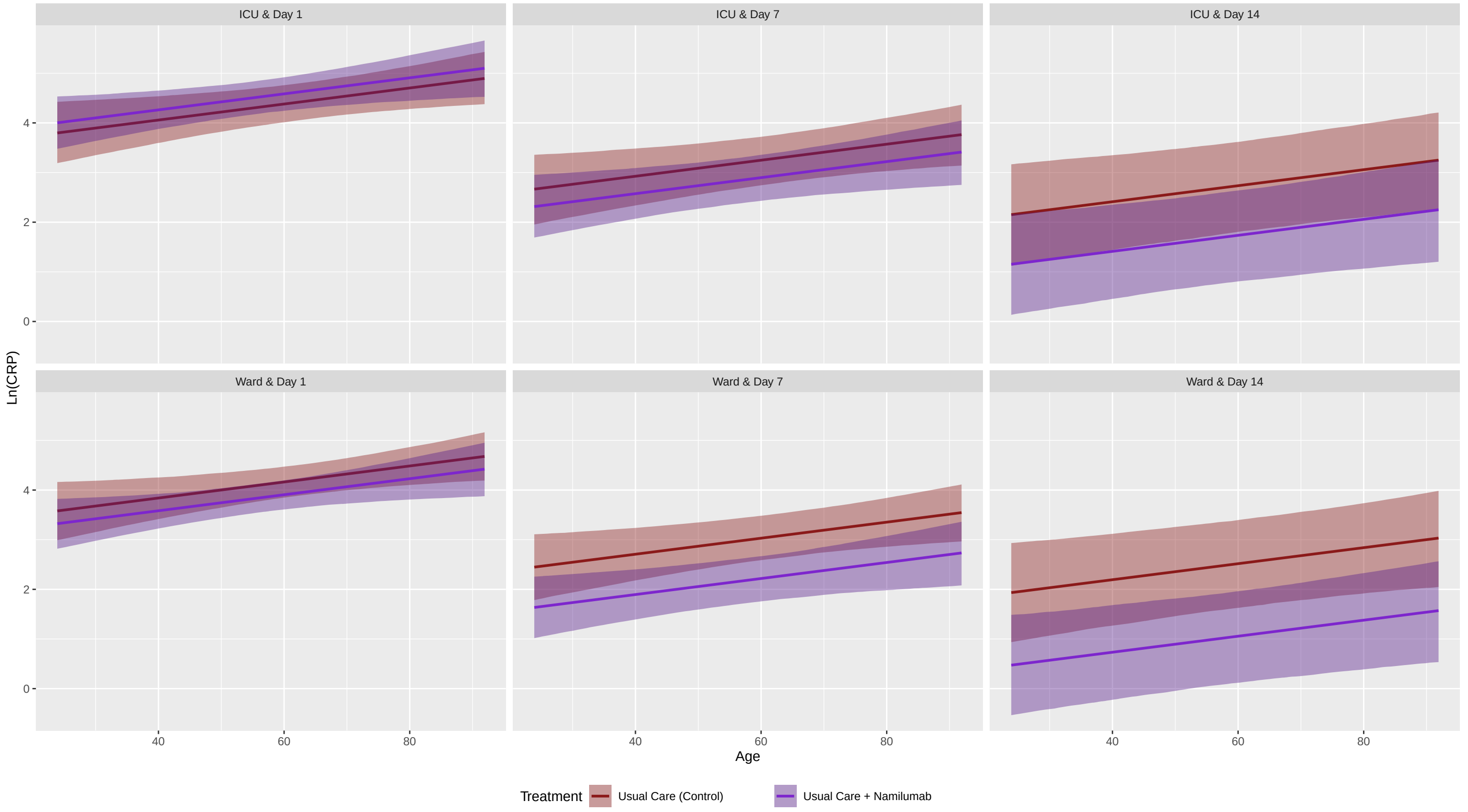


(a)

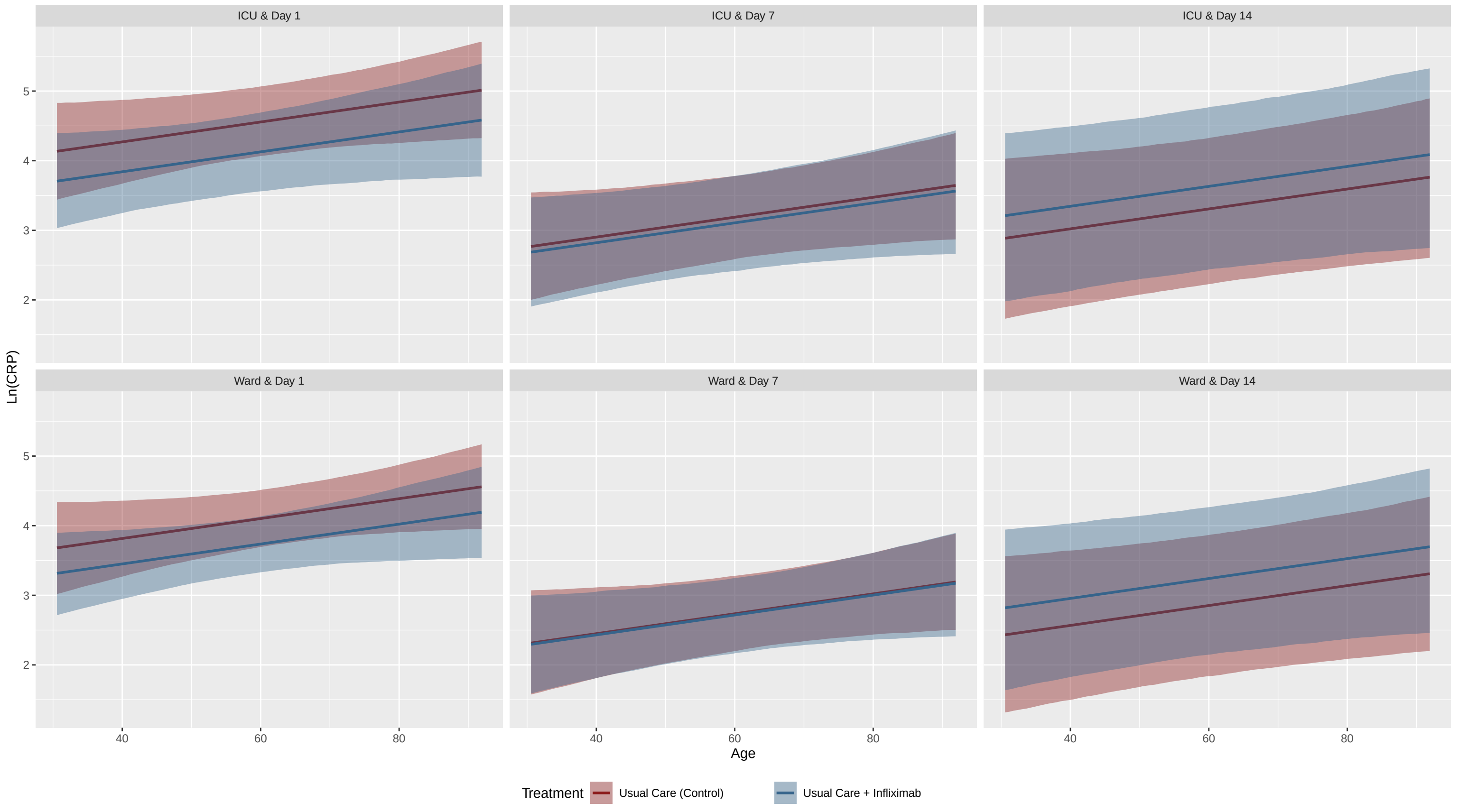


(b)

Figure 4: Supplementary Figure 4. Kaplan-Meier plots for time to 2 point improvement for whole population (A, D), ward (B, E) and ICU (C, F) for namilumab (A-C) and infliximab (D-F).

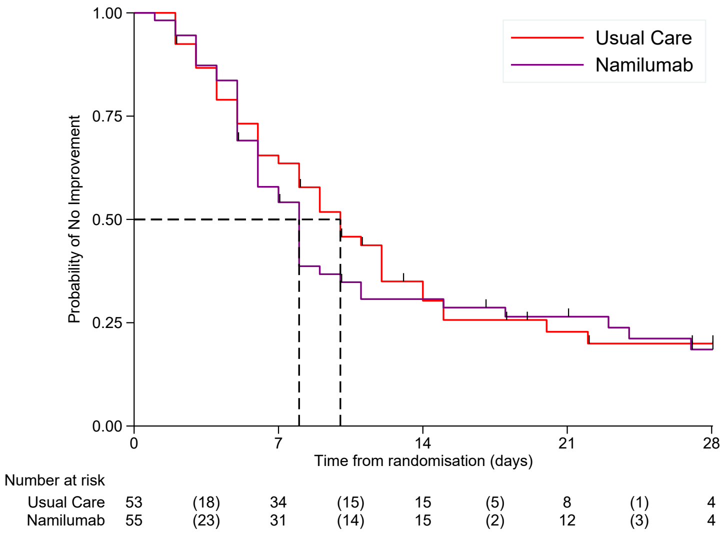


(a)

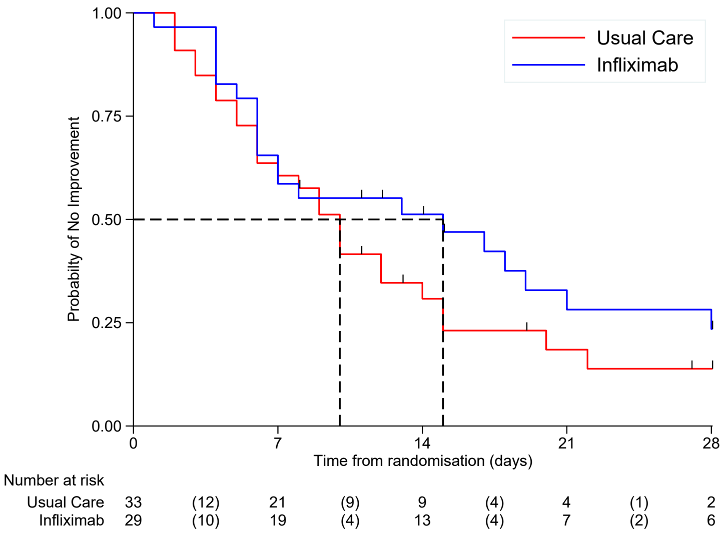


(b)

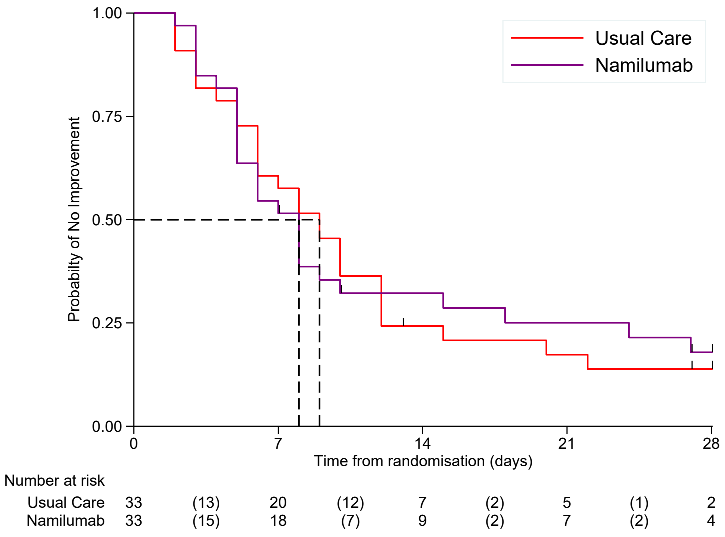


(c)

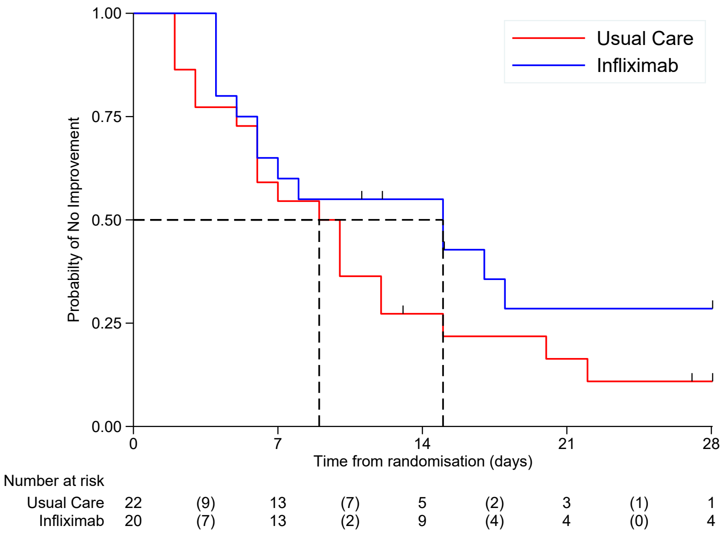


(d)

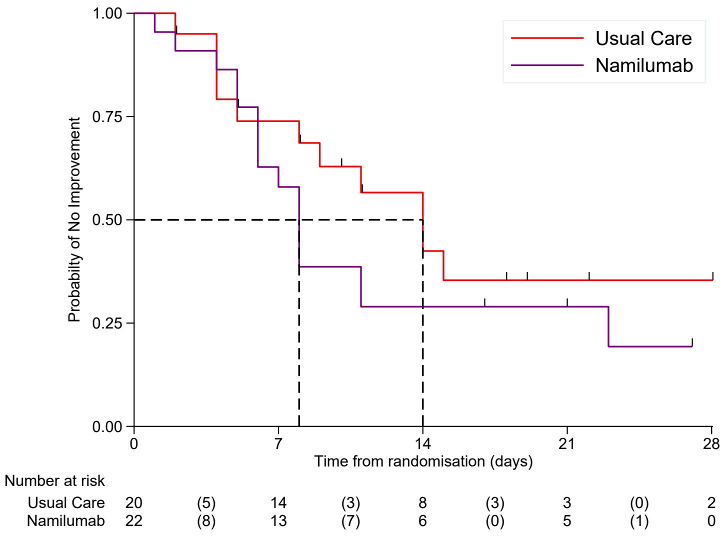


(e)

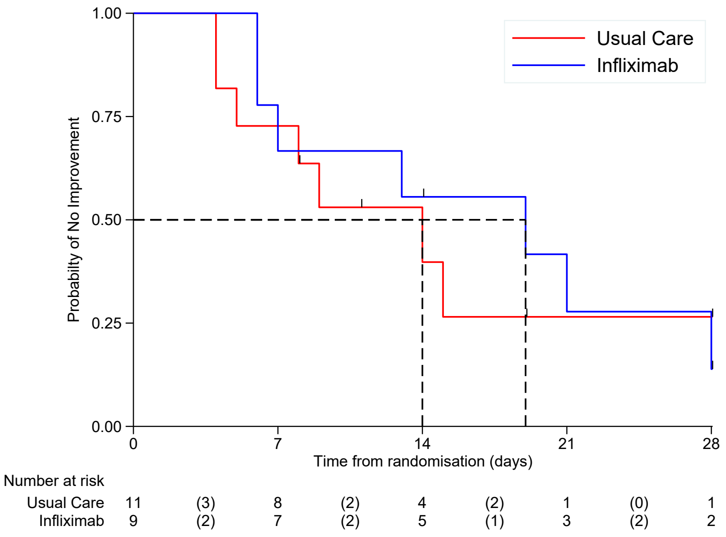


(f)

Figure 5: Supplementary Figure 5. Median oxygen saturation to fraction of inspired oxygen ratio (SF ratio) over time (days) for (A) namilumab (n=55 namilumab and n=54 usual care) and (B) infliximab (n=34 usual care and n=29 infliximab). Higher values indicate better oxygenation status.

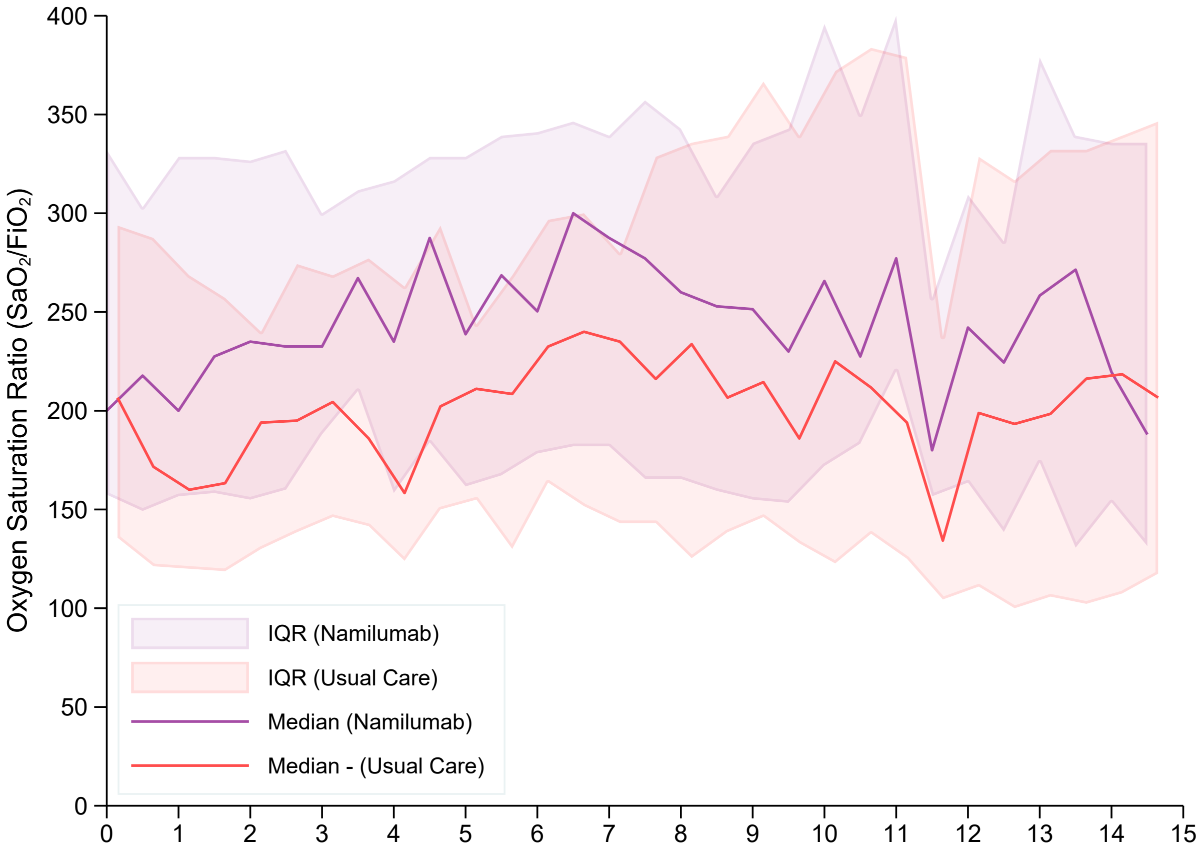


(a)

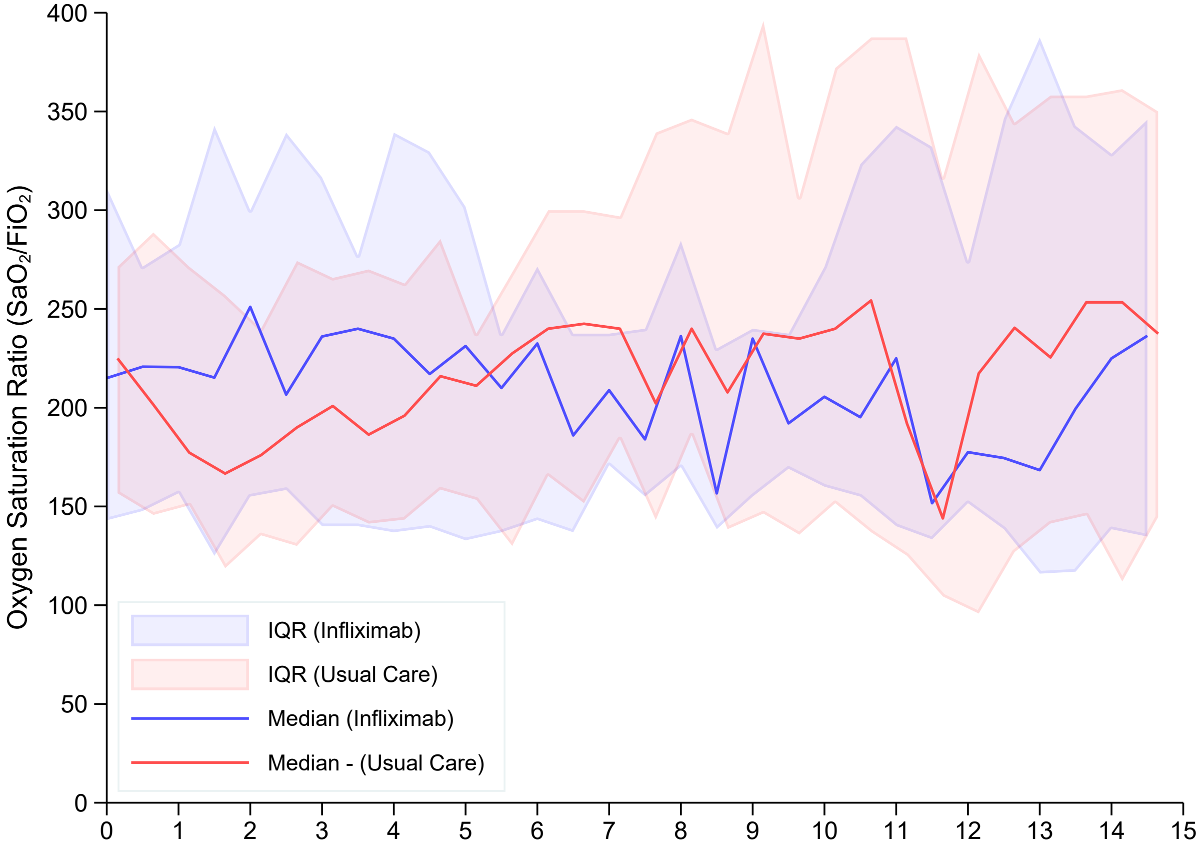
